# A retrieval-augmented generation large language model framework for accurate dementia identification from electronic health records

**DOI:** 10.64898/2026.01.24.26344477

**Authors:** Liqin Wang, Baoren Liu, Richard Yang, Ya-Wen Chuang, Hossein Estiri, Shawn Murphy, Li Zhou, Gad A. Marshall

## Abstract

**Objective:** Accurate and scalable dementia phenotyping from electronic health records (EHRs) is foundational for population-level research, risk prediction, and learning health system interventions. Traditional rule- and keyword-based approaches are limited by inconsistent documentation and inability to capture clinical nuance. We aim to develop and evaluate a framework that leverages large language models (LLMs) with retrieval-augmented generation (RAG) to overcome these limitations and improve dementia identification from real-world EHR data.

**Methods:** Using EHR data from the Mass General Brigham health system, we first assembled a cohort of adults with potential dementia based on diagnosis codes, problem lists, dementia-related medications, and free-text note mentions. A subset of candidate cases underwent detailed manual chart review to assign gold-standard dementia status. With this labeled sample, we implemented and compared three approaches for dementia ascertainment: (1) a rule-based classifier leveraging structured EHR data, (2) large language models (LLMs) applied to keyword-filtered clinical note excerpts, and (3) a RAG-based LLM framework that integrates retrieved, context-rich note snippets. Within each approach, we evaluated multiple configurations of embedding models, retrieval methods, LLMs, structured-data inclusion, and prompts to identify the best-performing classifier. Performance was assessed using standard classification metrics, including sensitivity, specificity, positive predictive value (PPV), and F1 score, and supplemented by qualitative error analyses to characterize common sources of false positives and false negatives across methods.

**Results:** The RAG-based classifier achieved the highest performance (F1=0.933, sensitivity=91.1%, PPV=95.5%) compared to rule-based (F1=0.823, sensitivity=81.1%, PPV=83.5%) and keyword-filtered LLM (F1=0.903, sensitivity=91.7%, PPV=88.6%). Including ICD codes alongside free text in the RAG-based LLM pipeline significantly reduced the PPV and modestly decreased F-1 score. Error analysis revealed that structured-code dependence contributed to false positives, whereas unrecognized contextual cues in notes drove false negatives.

**Conclusion:** A RAG-based LLM pipeline without structured ICD codes improved dementia ascertainment from EHR data compared with ICD-based rules and keyword-based filtering. This approach can enhance dementia case identification and support patient care, predictive modeling and risk analysis.

## INTRODUCTION

As the global population ages and the prevalence of dementia rises, the demand for early and accurate diagnosis of dementia has intensified, particularly given the emergence of the FDA-approved disease-modifying treatments for early Alzheimer’s disease (AD).^1–3^ Accurate ascertainment of dementia status in electronic health records (EHRs) is essential not only for clinical care but also for research using real-world data, including artificial intelligence (AI) models for risk stratification and disease prognosis, which in turn support efforts toward earlier detection and intervention.^4, 5^

Dementia is often underdiagnosed or inconsistently documented in EHRs, partly due to its gradual onset, shortage of specialists, insufficient time to assess patients, and variability in clinical documentation. Traditional methods for identifying dementia have primarily relied on structured EHR data, such as international classification of disease (ICD) diagnostic codes, medications and cognitive test results.^6–10^ While useful, these methods are limited by the variability in how dementia is recorded, with many details captured in unstructured clinical notes rather than in the standardized EHR fields. More recent machine learning models have demonstrated improved performance when combining structured and unstructured EHR data.^11–14^ However, most models are developed and evaluated within a single health system, and their performance can degrade substantially when applied to populations, care settings, or documentation practices that differ from the training data.^6, 15^

Large language models (LLMs), pre-trained on large, heterogeneous corpora, offer new opportunities to process free-text notes with greater contextual understanding and to generate interpretable reasoning beyond traditional natural language processing (NLP) or black-box machine learning models. They can often be applied “out of the box” using in-context prompting or with minimal find-tuning on local data, reducing the need for large amount of site-specific labeled data.^16^ Applying LLMs directly to clinical notes is impractical. Concatenating all notes typically exceeds model input limits, while processing notes individually prevents the model from capturing the patient’s longitudinal context and overall clinical picture. To address this, existing approaches often rely on condensing records through summarization or keyword-based filtering,^17, 18^ but these methods may omit clinically important details or introduce bias. Retrieval-augmented generation (RAG) has emerged as a promising strategy to address this challenge by embedding and indexing records, then retrieving only the most relevant segments for a given task.^19, 20^ This allows the model to consider longitudinal evidence without processing entire charts, thereby mitigating truncation and reducing noise. In this study, we developed and evaluated a RAG-based approach alongside rule-based and keyword-filtered LLM approaches for dementia ascertainment, using chart-reviewed data as the reference standard. We hypothesized that the RAG-based approach will outperform the other approaches.

## METHODS

### Clinical Setting

The study was conducted at Mass General Brigham (MGB), one of the largest integrated healthcare systems in the New England area. MGB comprises two founding hospitals (i.e., Massachusetts General Hospital, Brigham and Women’s Hospital), and various community and specialty hospitals. The study was approved by the MGB Institutional Review Board with waiver of informed consent from study participants owing to secondary use of EHR data.

### Data Source

MGB has been using EHR systems since the 1980s, providing over 40 years of longitudinal data to support research. The Research Patient Data Registry (RPDR) at MGB is a centralized repository that integrates clinical data across the health system, including demographics, encounters and diagnoses, laboratory tests and procedures, medications, patient-reported outcomes, and clinical notes.^21^ RPDR enables basic cohort identification through queries to structured fields and keyword searches of notes, as well as extraction of detailed EHR data and downstream analyses. EDW is a centralized repository at MGB that stores integrated data from various sources within an organization, used for reporting, analytics, and decision-making.

### Study Population

We searched MGB’s RPDR and EDW databases from January 1, 2010, to January 31, 2024, to identify a large cohort of patients with potential dementia. The search was conducted using a set of codes and keywords as described in **Table S1**. The EHR components searched included problem lists, encounters and diagnoses, flowsheets, medication prescriptions, and clinical notes. From the pool of potential dementia patients, we randomly selected 700 patients to form the initial study cohort for chart review. For this cohort, we retrieved structured and unstructured EHR data of the study cohort from RPDR from July 5, 1983, up until January 31, 2024.

Structured data included demographics, diagnoses, problem lists, and medications. The unstructured EHR data included progress notes, visit notes, discharge summaries and radiology reports. Patients without data from the RPDR were excluded. We then obtained ADRD labels via chart review. Of the study cohort with labels, we reserved 5% of the cases for algorithm development, such as prompt engineering, and the remaining for evaluation.

### Reference Standard for Dementia Classification

The study aimed to evaluate and compare multiple LLM-based pipelines to identify the most effective approach for determining the presence of dementia. The ground truth was established through a chart review of the sampled cohort. Two subject matter experts (Y.C. and L.W.) independently assessed the EHR data for each case up to January 31, 2024, utilizing EPIC Hyperspace^®^ and RPDR data to verify the presence of dementia. The finalized label of each case was confirmed through consensus between the two reviewers, or by a third reviewer (G.A.M.) in the event of a conflict. During the chart review process, we excluded patients who did not have medical records in the RPDR. Given that the algorithms relied on RPDR data only, we ensured that the patients who were labeled as positive for dementia had the evidence from RPDR data. Otherwise, the patients were labeled as negative for dementia.

### Dementia Classification Algorithms

We implemented and compared three approaches for dementia ascertainment (**Fig. 1**). First, the rule-based approach determined dementia status based on the number of relevant ICD codes and served as the baseline. Second, the keyword-filtered LLM approach combined ICD codes and expert-defined keywords to extract relevant sentences, which were then passed to LLMs for classification. Third, the RAG-based LLM approach used semantic retrieval to extract clinically relevant text chunks and passed into LLMs for classification. Details of the three approaches are described below.

**Fig. 1.**
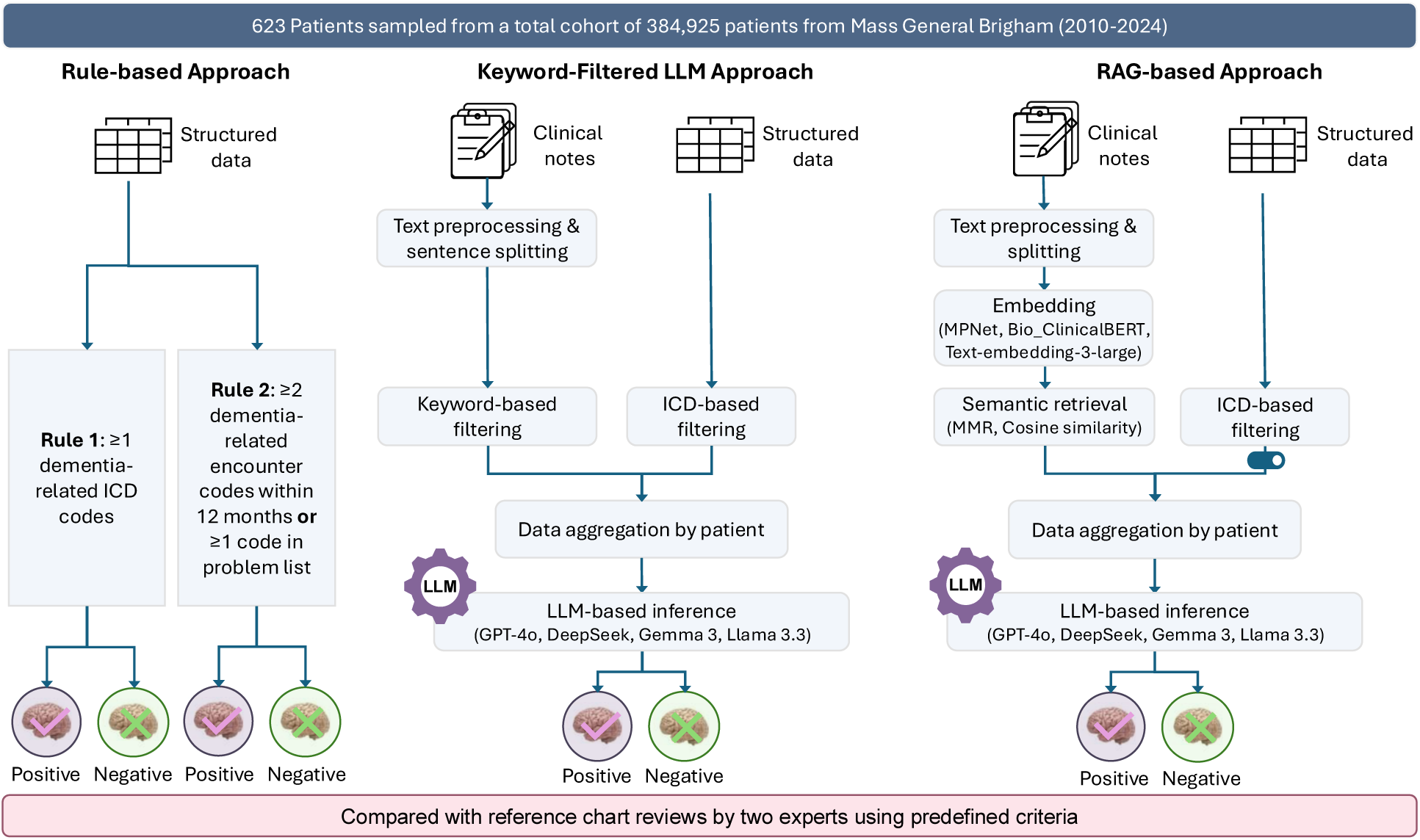
Overview of rule-based, keyword-filtered LLM, and RAG-based approaches for dementia classification. Three approaches were developed and applied to 623 patients sampled from a total cohort of 384,925 patients at Mass General Brigham (2010–2024). The rule-based approach identified dementia using structured data through (Rule 1) ≥ 1 dementia-related ICD code or (Rule 2) ≥ 2 encounter codes within 12 months or ≥ 1 code in the problem list. The keyword-filtered LLM approach used expert-defined keywords and ICD codes to extract dementia-related text from clinical notes for large language model (LLM) inference. The RAG-based LLM approach incorporated text preprocessing, embedding generation, and semantic retrieval to enhance context-aware information extraction prior to LLM inference. All classifier outputs were compared with expert-adjudicated reference standards.

#### Rule-based approach

We applied a rule-based approach using dementia-related ICD-9-CM and ICD-10-CM codes (**Table S2**). In Rule 1, patients were classified as having dementia if they had any such code in the problem list or encounter diagnoses. In Rule 2, patients were classified as having dementia if they had ≥2 dementia-related encounter diagnoses within 12 months or ≥1 such code in the problem list.

#### Keyword/ICD-based LLM approach

The keyword-filtered LLM approach streamlined dementia classification by filtering each patient’s records to retain only relevant evidence before applying LLM. It consisted of four steps: information retrieval, data aggregation, prompt development, and dementia classification.

##### Information retrieval

Processing lengthy texts increases computational cost and latency for transformer-based LLMs whose memory and processing demands scale with input length. Because dementia-related information typically appears in only a small portion of notes, we extracted relevant content prior to LLM input. Using a predefined list of keywords (**Table S3**) consolidated from two prior studies and manually curated by L.W. and G.A.M,^22, 23^ we identified sentences related to cognitive decline or dementia. Clinical text was segmented with Stanza;^24^ sentences containing at least one keyword were retained using partial matching (exact token matching for abbreviations). To avoid redundancy from copy-paste content, we excluded sentences repeated in prior notes. For context, we included the sentences immediately before and after each selected sentences to form short paragraphs. Structured data like problem lists and diagnoses were filtered using dementia-related ICD-9/10-CM codes (**Table S2**).

##### Data aggregation

For each patient, filtered information was ordered chronologically. Structured data (diagnoses and problem lists) and unstructured data (clinical notes) from the same day were concatenated into a standardized daily template (**Table S4**). Only data elements present for a given day were included; if no diagnosis, problem list, or clinical notes were present for that day, the corresponding field was omitted. The resulting daily summaries were then aggregated across time to form a longitudinal patient record.

##### Prompt development

We developed and tested prompts using 33 randomly selected patients from the reference dataset, which were excluded from evaluation. A zero-shot strategy was applied to assess pre-trained models for dementia identification. The initial prompt defined model’s role, task, key instructions, requiring a binary ’YES’ or ’NO’ response. Prompts were iteratively refined mainly through insights from error analysis. As summarized in **Table S5**, each setup consisted of a system prompt (defining roles and output requirements) and a user prompt (containing patient data and a classification query). We evaluated three versions of the classification query, which differed in instructional style and level of specificity. Specifically, Prompt 1 (P1) incorporated explicit interpretation guidelines for cognitive test scores (MMSE, MoCA, and Mini-Cog), which are frequently mentioned in clinical notes. Prompt 2 (P2) provided a high-level framework, emphasizing what to prioritize and what to disregard when identifying dementia. Prompt 3 (P3) introduced a structured, stepwise process that required supporting evidence at each stage, making the decision pathway more detailed and prescriptive. We evaluated these three prompts based on the best-performing LLM identified in the previous step and compared their classification performance.

##### Dementia classification

For patients with filtered data, we deployed both open-source and commercial LLMs. Open-source models included Gemma 3 27B (Google), Llama3.3 70B (Meta), and DeepSeek-R1 70B (DeepSeek), ran locally on a dual NVIDIA L4 GPU server via Ollama’s Python library (see **Table S6**). For commercial deployment, we used GPT-4o via the Microsoft Azure platform, which is Health Insurance Portability and Accountability Act (HIPAA) compliant. All models were configured with a temperature of 0 to ensure consistent and precise outputs. Each LLM produced a binary dementia classification along with explanatory text.

Patients with no relevant EHR data after keyword/ICD filtering were classified as negative dementia.

#### RAG-based LLM approach

Unlike the keyword-filtered LLM classification, which relies on predefined terms to select relevant EHR content, the RAG-based method dynamically retrieves information from the full EHR corpus using retrieval-augmented generation. Retrieved text is then passed to the LLM for classification. The RAG pipeline consists of four components: document segmentation, embedding, retrieval, and response generation. By grounding the LLM’s output in retrieved evidence, this approach enables more context-aware and adaptive dementia classification. As summarized in **Table 1**, we conducted 6 experiments (A-F) encompassing 12 unique RAG pipelines. Each experiment isolated the effect of a single component (e.g., embedding model in Experiment A) while holding others constant. Given the large number of possible combinations across embedding models, retrieval methods, LLMs, structured-data inclusion, and prompts for response generation, we adopted a staged design: in each experiment, the first row represented the baseline pipeline carried over from the previous experiment, either the best-performing or retained pipeline if no subsequent improvement was observed, serving as the reference for comparison.

**Table 1.**
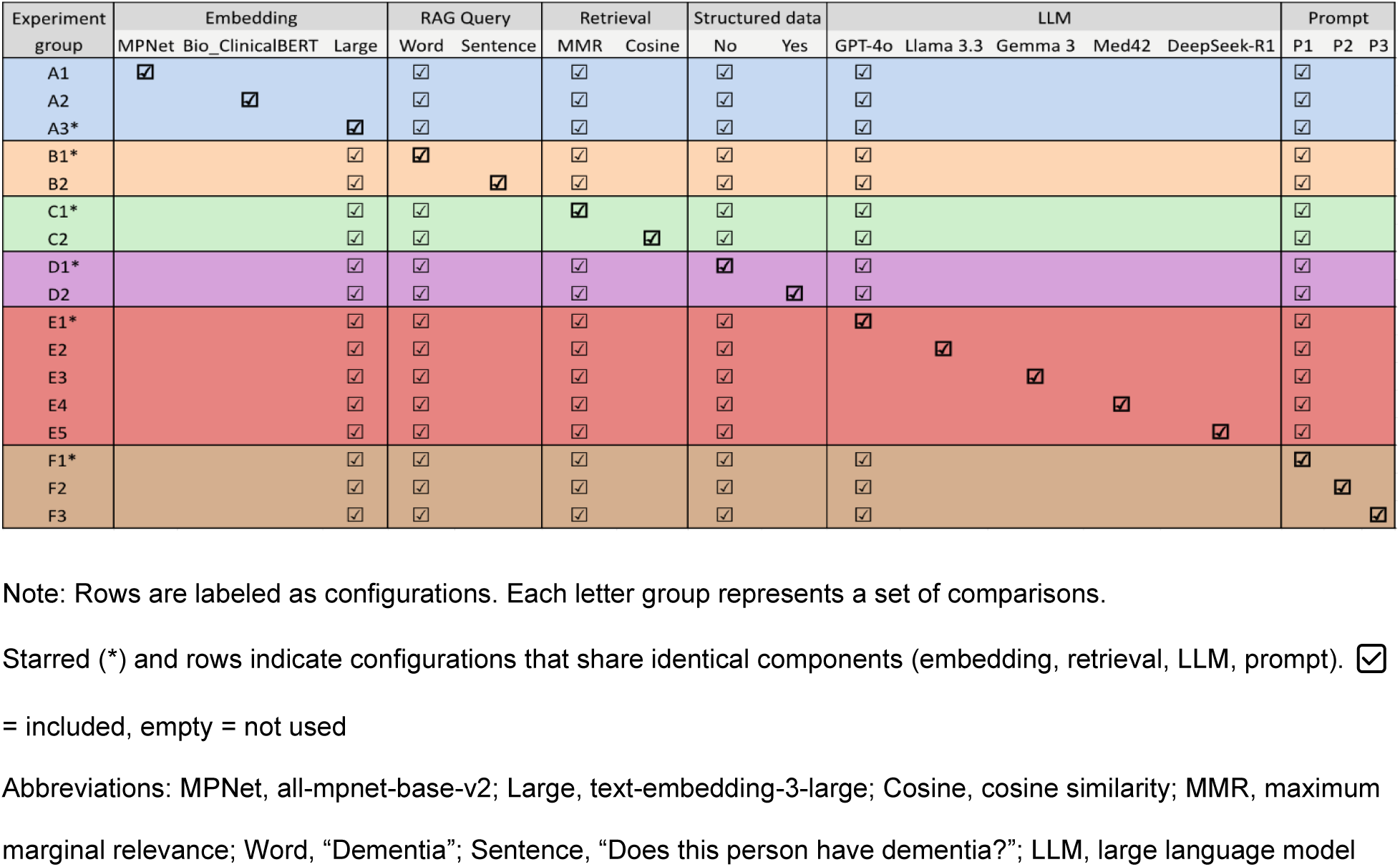
Detailed comparison of various settings in a retrieval-augmented generation (RAG)-based large language model (LLM) framework.

##### Document segmentation

Clinical notes can be lengthy and contain many sections that are not relevant to cognitive decline or dementia. We performed section-based splitting, which uses a rule-based NLP approach to segment clinical notes into sentences and then iteratively searches for section headers using regular expressions and a predefined dictionary. When a new section header is detected, subsequent sentences are grouped under that section until the next header is found. Each document was split into one or more sections.

##### Text embedding

After splitting the documents into chunks, we applied a bi-encoder technique to embed text chunks into semantic vector representations. Embeddings were generated using a mix of general and domain-specific models, including all-mpnet-base-v2, text-embedding-3-large, and the medically focused BioClinicalBERT (**Table S7**). These embeddings were stored in ChromaDB, a vector database, to prepare for retrieval. We evaluated these models in the dementia classification pipeline (Experiment A in **Table 1**)

##### Document retrieval

To retrieve dementia-related information, we used the same embedding model to generate a vector for the query phrase (e.g., "Dementia" or "Does the patient have dementia?") and tested which query phrase yielded better performance (Experiment B, **Table 1**). A distance function, cosine similarity, was then applied to identify the top-k closest text chunks (k=30) for individual patients. Due to frequent duplication in clinical notes from copy-pasting, cosine similarity often surfaced redundant chunks. To address this, we implemented MMR, which balances both relevance and diversity in retrieval. We compared cosine similarity and MMR performance as described in Experiment D (**Table 1**). For structured data, similar as the keyword-filtered LLM approach, we used predefined ICD-9/10-CM codes to filter relevant information from problem list and diagnoses (see **Table S2**).

##### Dementia classification

For LLM-based classification, each of the top-k retrieved text chunks was concatenated with corresponding metadata, such as report number, date, and type, and then ordered chronologically by report date. Instructions and questions were appended after patient information. The structured data, such as problem list and diagnosis codes, might be essential for understanding health conditions; however, some diagnosis codes may reflect billing activities rather than actual conditions. To evaluate the impact of incorporating structured data on dementia classification, we compared prompts with and without feeding this data (Experiment D, **Table 1**). In both with and without structured data pipelines, patients with only structured data were classified using the filtered structured data alone. Additionally, we deployed both open-source and commercial LLMs and compared their performance while controlling other pipeline components (Experiment E, **Table 1**). Lastly, we evaluated three versions of prompts in the RAG-based pipeline, which matched those used in the keyword-filtered LLM pipeline (Experiment F, **Table 1**).

## Statistical Analysis

We assessed the performance of the rule-based approach, keyword-filtered LLM classification, and RAG-based LLM classification by calculating the positive predictive value (PPV), sensitivity, specificity, accuracy, and F1 score. We optimized the pipeline for the best F1 score because it is a balanced metric between PPV and sensitivity. To gain a comprehensive understanding of each algorithm’s strengths and weaknesses, we conducted a detailed error analysis focusing on false positives (FPs) and false negatives (FNs) for the best-performing pipelines from the three approaches: rule-based, keyword-filtered LLM, and RAG-based LLM. We calculated the error composition of each classifier among the 623 patients compared with the reference standard. In addition, we examined the overlap of FP and FN cases across the three approaches to assess the frequency of errors and conducted a detailed review of grouped cases to identify the underlying causes of these errors.

## RESULTS

We identified 384,925 patients aged ≥50 years with potential dementia from the Mass General Brigham (MGB) EHR databases between January 1, 2010 to January 31, 2024. From this sample, 700 patients were randomly selected to establish the reference cohort. Of these, 44 (6.3%) were labeled as unknown for dementia due to insufficient records. The remaining cases included 33 patients (5%) for prompt refinement and 623 for evaluating the three pipelines (**Fig. 1**). Among the 623 patients, 366 (58.7%) were female; the mean aged was 73.9 years (based on the last available records in the MGB system or study end date, whichever occurred first), and 169 (27.1%) were positive for AD and related dementias (ADRD) (**Table 2**).

**Table 2.**
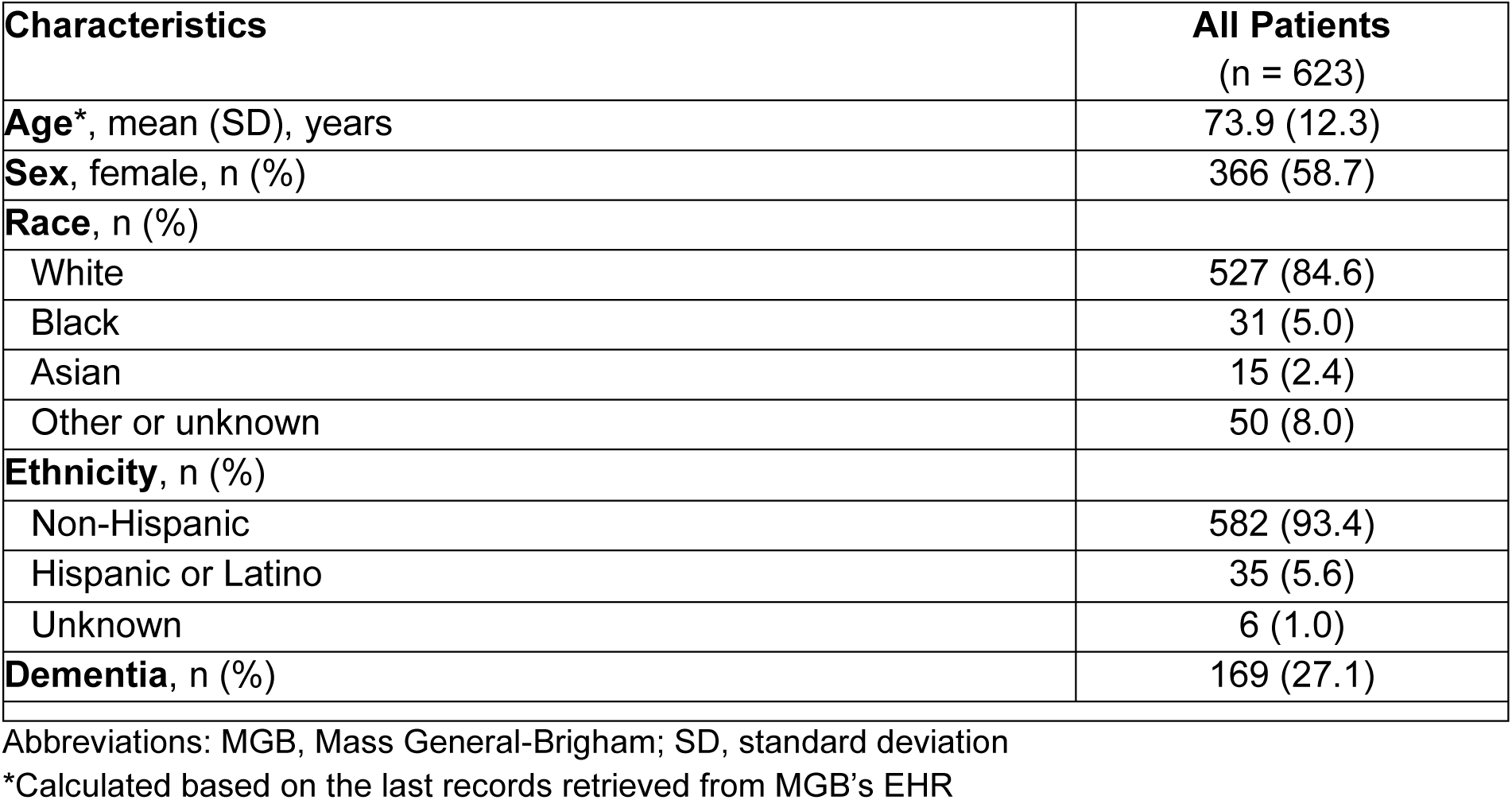
Characteristics of 623 Patients from Mass General Brigham’s Electronic Health Records.

| **Characteristics** | **All Patients**<br>**(n = 623)** |
| --- | --- |
| **Age\***, mean (SD), years | 73.9 (12.3) |
| **Sex**, female, n (%) | 366 (58.7) |
| **Race**, n (%) |  |
| White | 527 (84.6) |
| Black | 31 (5.0) |
| Asian | 15 (2.4) |
| Other or unknown | 50 (8.0) |
| **Ethnicity**, n (%) |  |
| Non-Hispanic | 582 (93.4) |
| Hispanic or Latino | 35 (5.6) |
| Unknown | 6 (1.0) |
| **Dementia**, n (%) | 169 (27.1) |

Abbreviations: MGB, Mass General-Brigham; SD, standard deviation
\*Calculated based on the last records retrieved from MGB’s EHR

### Rule-based approach

Between the two rule-based approaches, the Rule 1 classifier (R1) achieved better overall performance (F1=0.823) (**Table 3**). Among 169 patients with dementia, 81.1% (n=137) had at least one dementia-related ICD code, while 18.9% (n=32) had none. Of 164 patients with dementia-related ICD codes in either diagnosis or problem list, 16.5% (n=27) patients did not have dementia, suggesting possible coding for billing purposes. Compared to Rule 1, Rule 2 slightly increased the positive predictive value (PPV) from 83.5% to 87.6% but reduced the sensitivity from 81.1% to 58.6%.

**Table 3.**
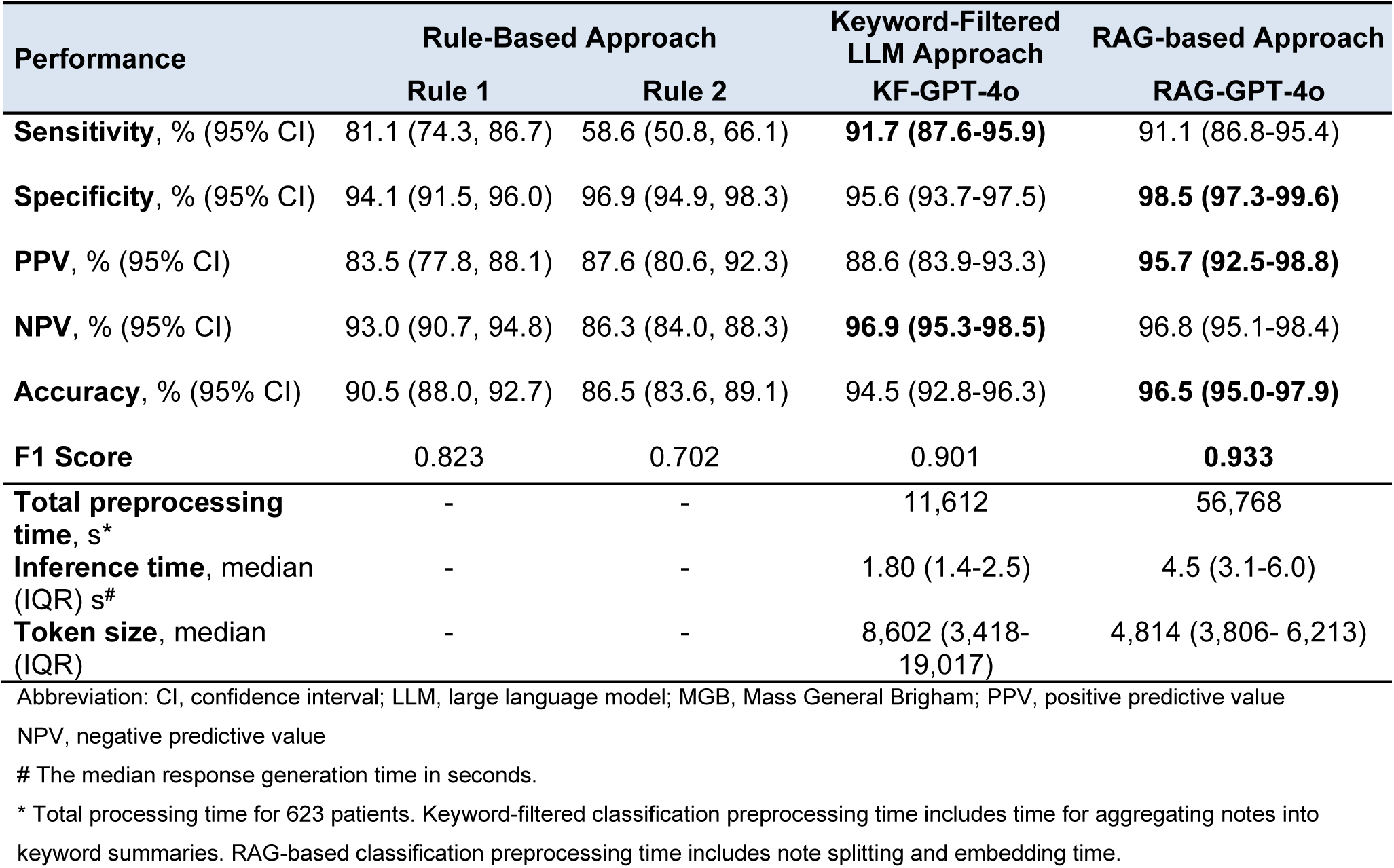
Performance of the best classifiers of the keyword-filtered LLM and RAG-based approaches compared with two rule-based algorithms.

| Performance | Rule-Based Approach |  | Keyword-Filtered LLM Approach | RAG-based Approach |
| --- | --- | --- | --- | --- |
|  | Rule 1 | Rule 2 | KF-GPT-4o | RAG-GPT-4o |
| **Sensitivity**, % (95% CI) | 81.1 (74.3, 86.7) | 58.6 (50.8, 66.1) | **91.7 (87.6-95.9)** | 91.1 (86.8-95.4) |
| **Specificity**, % (95% CI) | 94.1 (91.5, 96.0) | 96.9 (94.9, 98.3) | 95.6 (93.7-97.5) | **98.5 (97.3-99.6)** |
| **PPV**, % (95% CI) | 83.5 (77.8, 88.1) | 87.6 (80.6, 92.3) | 88.6 (83.9-93.3) | **95.7 (92.5-98.8)** |
| **NPV**, % (95% CI) | 93.0 (90.7, 94.8) | 86.3 (84.0, 88.3) | **96.9 (95.3-98.5)** | 96.8 (95.1-98.4) |
| **Accuracy**, % (95% CI) | 90.5 (88.0, 92.7) | 86.5 (83.6, 89.1) | 94.5 (92.8-96.3) | **96.5 (95.0-97.9)** |
| **F1 Score** | 0.823 | 0.702 | 0.901 | **0.933** |
| **Total preprocessing time**, s* | - | - | 11,612 | 56,768 |
| **Inference time**, median (IQR) s<sup>#</sup> | - | - | 1.80 (1.4-2.5) | 4.5 (3.1-6.0) |
| **Token size**, median (IQR) | - | - | 8,602 (3,418-19,017) | 4,814 (3,806- 6,213) |

Abbreviation: CI, confidence interval; LLM, large language model; MGB, Mass General Brigham; PPV, positive predictive value
NPV, negative predictive value

### Keyword-filtered LLM approach

**Table S8** summarizes the performance of four LLMs evaluated with prompt P1 and three prompt configurations (P1-P3) tested using GPT-4o (see **Table S5** for the prompts). Among four models, GPT-4o achieved the best performance (accuracy = 94.5%, F1 = 0.901), substantially outperforming the open-source models (DeepSeek-R1: accuracy = 79.2%, F1=0.409; Llama-3.3: accuracy = 79.9%, F1=0.444; Gemma-3: accuracy = 45.9%, F1 = 0.499). Across the three prompts with GPT-4o, performance was consistent (F1 = 0.896-0.901), with P1 showing a slight advance. The keyword-filtered GPT-4o pipeline with P1 (KF-GPT-4o) was selected as the best-performing configuration.

### RAG-based approach

**Table 1** summarizes six experiments: Experiment A evaluated three embedding models; Experiments B-D tested retrieval components; and Experiments E-F evaluated LLMs and prompts for inference. **Table 4** shows the performance of 12 RAG-based pipelines.

- Experiment A compared three embedding models: all-mpnet-base-v2 (F1=0.914), Bio_ClinicalBERT (F1 = 0.874), and text-embedding-3-large (F1=0.923). The latter achieved the best performance by a small margin.
- Experiment B examined retrieval query phrasing. A single-word query (“Dementia”) achieved an F1 score of 0.923, compared with 0.924 for the full-sentence query (“Does this patient have dementia?”), while a negligible difference (<0.001) likely due to rounding. Subsequent experiments therefore used the single-word query.
- Experiment C compared two retrieval functions in RAG: maximal marginal relevance (MMR) achieved an F1 score of 0.923 outperformed cosine similarity (F1 = 0.913), confirming MMR as the superior method.
- Experiment D assessed the impact of incorporating structured data (i.e., ICD codes from problem list and encounter diagnoses) into the classification pipeline. Without structured data, the pipeline achieved a PPV of 98.0% and an F1 score of 0.923; adding structured data significantly reduced PPV to 88.9% as well as dropping F1 to 0.912, suggesting potential noise or misleading signals from structured fields.
- Experiment E compared five LLMs. GPT-4o achieved the highest performance (F1 = 0.923), followed by Med42 (0.868), while Gemma-3, Llama-3.3, and DeepSeek-R1 performed substantially worse (0.789, 0.764, and 0.755, respectively). These results indicate the model choice strongly impacts pipeline performance, with GPT-4o clearly outperforming the others.
- Experiment F compared three prompts. P1, P2, and P3 achieved F1 scores of 0.923, 0.922, and 0.933, respectively, indicating that P3’s step-by-step reasoning improved accuracy. However, inference time increased with prompt complexity: P1 was fastest (mean [SD]: 2.80 [1.04] s per patient), followed by P2 (3.24 [1.34] s) and P3 (4.74 [1.98] s), reflecting the added processing required for structured reasoning.

**Table 4.**
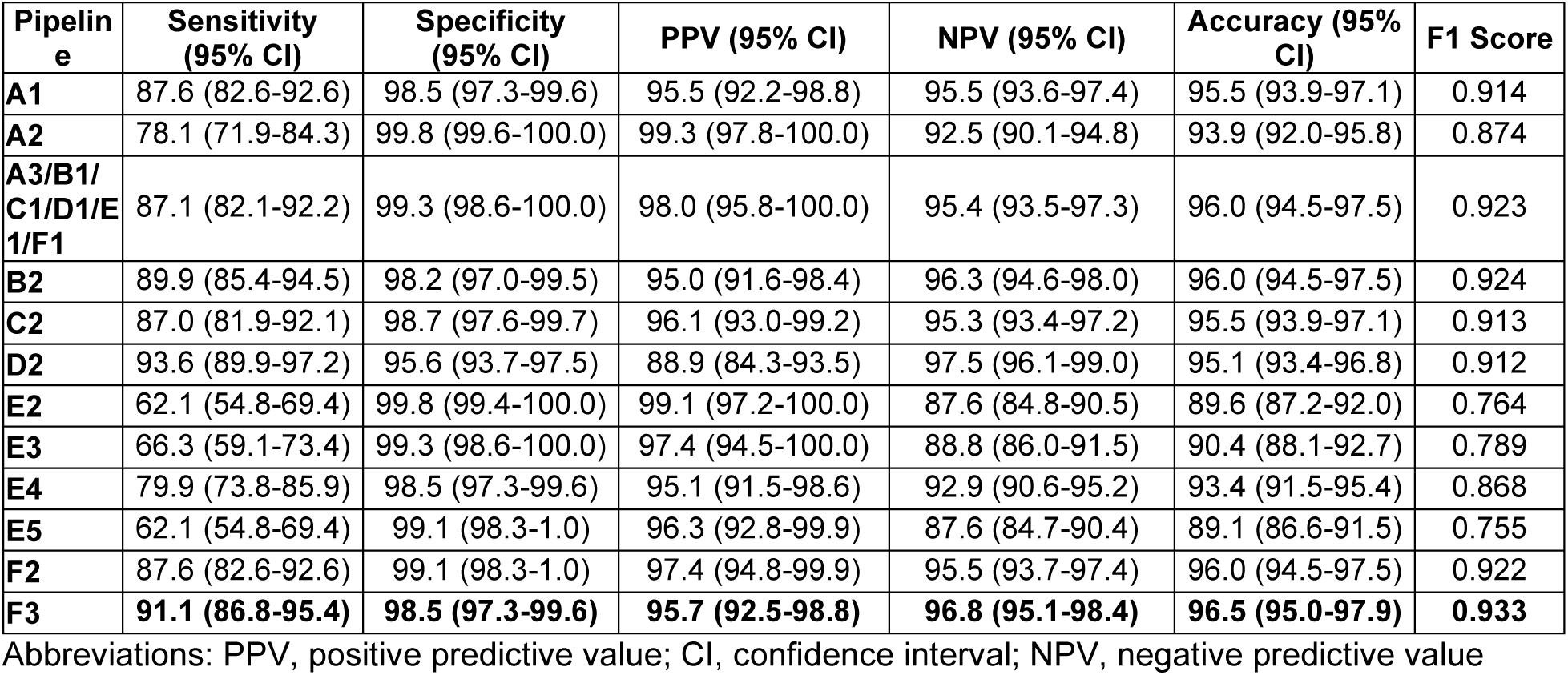
Performance of retrieval-augment generation (RAG)-based large language model (LLM) pipelines.

Across all experiments, the RAG-based GPT-4o (RAG-GPT-4o) classifier, using text-embedding-3-large embedding model, MMR-based retrieval with the single-word query, excluding structured data, and applying the structured prompt P3, achieved the best overall performance among the 12 pipelines.

Comparing the best classifiers from three approaches, RAG-GPT-4o (F1=0.933) outperformed KF-GPT-4o (F1=0.901) and R1 (F1=0.823) (**Table 3**). Notably, although KF-GPT-4o produced more tokens for LLM classification, the inference time and total processing time is much shorter than RAG-GPT-4o.

### Error Analysis

**Fig. 2** shows the confusion matrices of the best-performing classifiers from the three approaches: R1, KF-GPT-4o, and RAG-GPT-4o. Compared with R1, the KF-GPT-4o reduced false positives from 27 (4.3%) to 18 (2.9%) and false negatives from 32 (5.1%) to 15 (2.4%). RAG-GPT-4o produced the fewest false positives (7 [1.1%]) while maintaining similarly low false negatives (15 [2.4%]). These results indicate that both LLM-based classifiers (KF-GPT-4o and RAG-GPT-4o) substantially reduced classification errors, with RAG-GPT-4o showing the lowest false-positive rate. **Fig. 3** summarizes the overlap of false positive and false negative cases across the three classifiers.

**Fig. 2.**
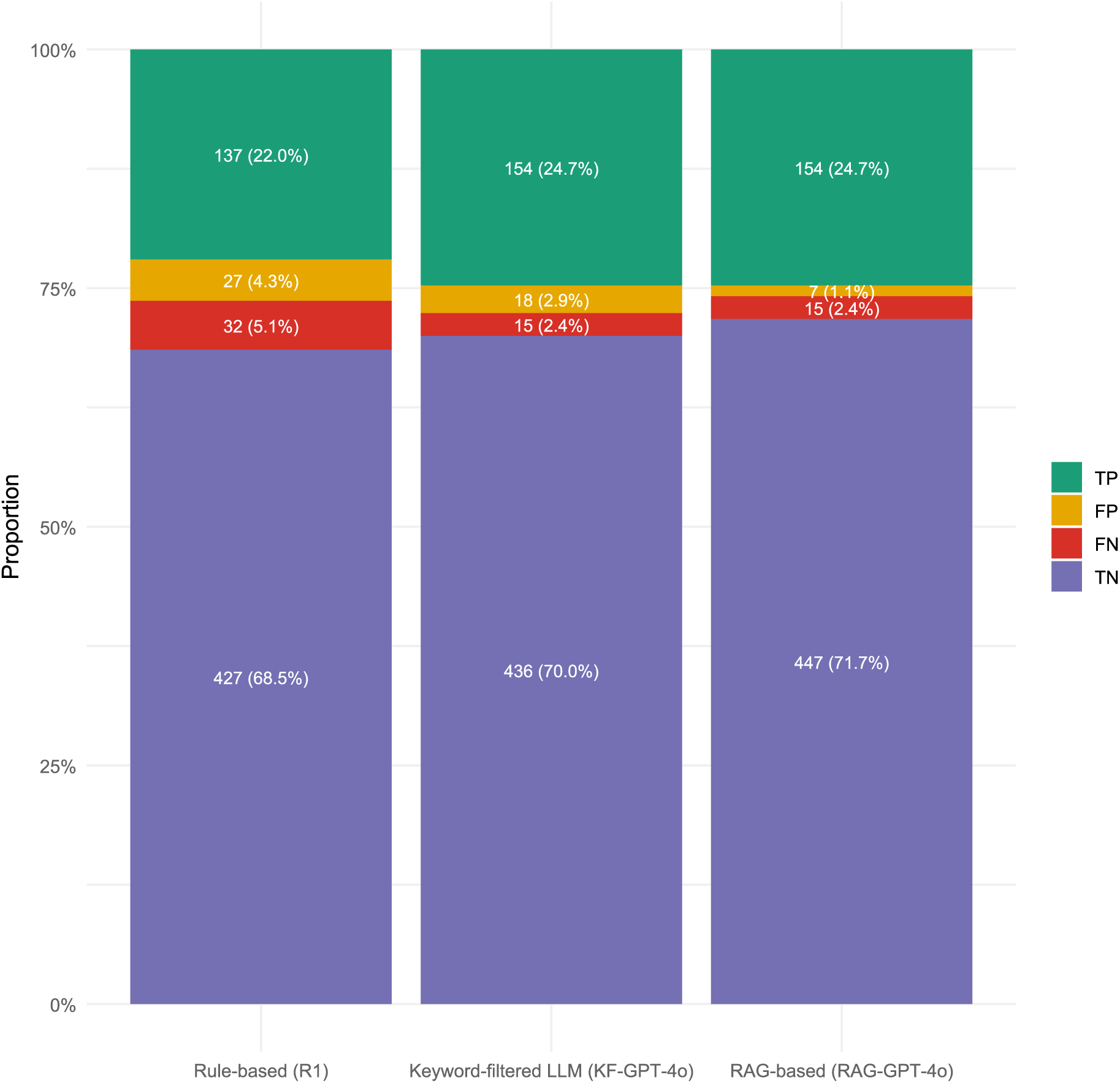
Error composition by classifier (counts and proportions).

**Fig. 3.**
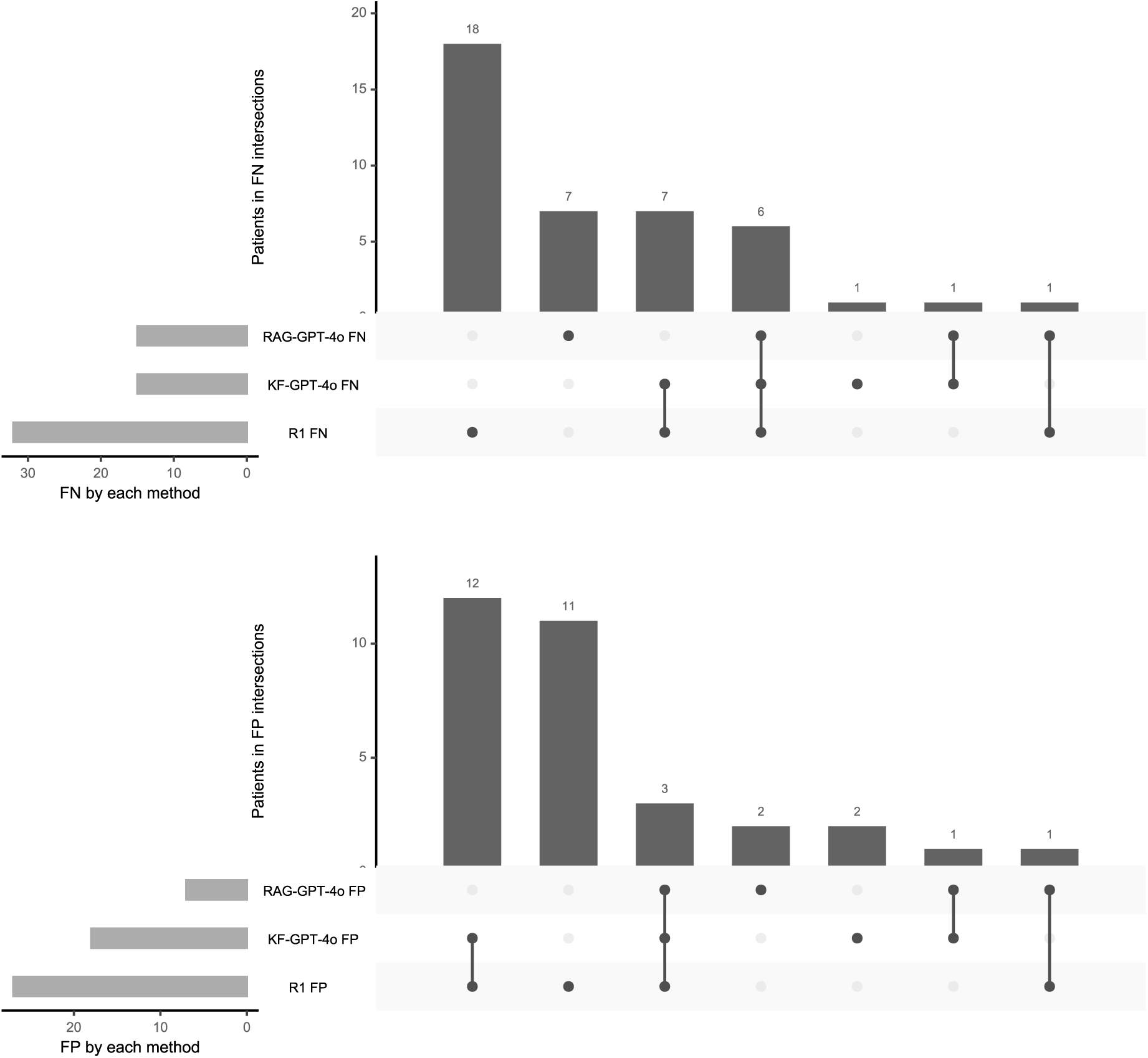
Overlap of false positive and false negative cases across dementia classification classifiers. (A) False negative (FN) cases where true dementia cases were missed. (B) False positive (FP) cases where patients were incorrectly classified as having dementia. Bars represent the number of patients in each intersection across the three pipelines: R1, KF-GPT-4o, and RAG-GPT-4o. Connected dots indicate which approaches contributed to each intersection.

#### False negatives

We examined overlaps in FN cases across the three classifiers. R1 misclassified 32 (18.9% of 169) dementia cases, 18 of which were unique to this classifier. All of them were due to missing dementia-related ICD codes in structured fields, despite clear evidence of dementia in the notes. The KF-GPT-4o produced 15 FNs, 13 overlapping with R1, indicating the absence of dementia-related ICD codes. Among these, 2 were caused by relevant information being truncated due to LLM input limits, 9 by the LLM overlooking available information, and 4 by differences in reasoning compared with human judgment, despite full context. RAG-GPT-4o produced 15 FNs, including 7 unique cases and 6 overlapping with both R1 and KF-GPT-4o. Among these, 5 were caused by missing key information during retrieval, 2 from the LLM not using key information included in the prompt, and 7 from differences in reasoning compared with human reviewers despite having and using relevant information.

Notably, only one FN case had evidence solely in the structured fields (i.e., problem list), and this information was excluded from the RAG-based pipeline.

#### False positives

The rule-based approach produced the most FPs (27/623, 4.3%), largely due to ICD codes entered for rule-out diagnosis and billing purpose rather than confirmed dementia. Of these 27, 15 were also flagged positives by the KF-GPT-4o because dementia-related ICD codes were concatenated to the clinical notes and misled the LLM. The remaining 12 were classified as negative, as the appended diagnosis codes appended after the lengthy concatenated notes exceeded GPT-4o’s input limit, and were truncated before reaching the model. RAG-GPT-4o produced 7 (1.1%) FPs, primarily due to diagnostic uncertainty between MCI and mild dementia. For example, some cases had low MoCA scores attributable to mood disorder or hearing loss, or notes indicated a history of dementia at baseline despite normal MoCA results.

## DISCUSSION

In this study, we evaluated the feasibility and performance of LLMs for ascertaining dementia status from EHR data at a major U.S. academic health system. Leveraging a system-wide search across the enterprise data warehouse (EDW) databases, we assembled a large candidate cohort and established reference-standard labels through chart review. We developed and compared three approaches (i.e., rule-based, keyword-filtered LLM, and RAG-based LLM), and within each approach, evaluated multiple configurations. LLM-based methods substantially outperformed traditional rule-based algorithms, with the RAG-based GPT-4o pipeline (text-embedding-3-large, MMR retrieval with a single-word query, no structured ICD data, prompt P3, GPT-4o) achieving the highest accuracy. These findings highlight the potential of LLMs, especially with retrieval augmentation, to enhance dementia phenotyping in real-world data and support epidemiologic, health services, and AI-driven prognosis research.

The traditional rule-based approach, though straightforward, demonstrated limitations in both sensitivity and PPV. About one in five dementia patients lacked dementia ICD codes in structured fields such as problem lists and diagnoses, indicating under-documentation in EHRs. Moreover, some patients with dementia-related ICD codes did not have true dementia, further underscoring the limitations of relying solely on coded data. The best-performing RAG-based LLM classifier overcame challenges that the rule-based classifier could not, particularly cases missing ICD codes but showing clear indications of dementia in clinical notes, and cases where codes reflected billing or rule-out diagnoses. Enabling the screening of the EHR databases with such a classifier could enhance documentation accuracy, strengthening real-world ADRD research and patient care.

Notably, incorporating structured ICD data into LLM pipelines reduced precision and overall performance. A RAG-based pipeline with structured data showed decreased PPV compared to the pipeline without structured data. Encounter diagnoses are often unreliable due to billing, historical carryover, or inaccurate documentation, and feeding them directly into the LLM can introduce misleading signals. These findings underscore the need to revisit structured data with improved normalization, integration, and weighting strategies to reduce noise while preserving clinical value, for example, selectively including validated sources, such as confirmed problem lists.

Compared with the KF-GPT-4o classifier, the RAG-GPT-4o classifier achieved superior performance in retrieving relevant information. Keyword-based extraction often produced overly long notes due to duplicated, copy-pasted content and unweighted keyword matches. In contrast, the RAG-based approach retrieved the top-n most relevant sections through a targeted semantic search, with maximal marginal relevance further improving diversity and reducing redundancy. Although the RAG pipeline occasionally missed relevant text, resulting in a few false negatives, the keyword-based method was more prone to truncating or overlooking recent information, particularly in patients with lengthy notes.

Given that the RAG pipeline consists of multiple configurable components that could yield thousands of combinations, our staged experimental design enabled systematic evaluation of key elements, including embedding models, retrieval strategies, structured data inclusion, LLM selection, and prompt formulation, while avoiding unnecessary runs. This methodical approach efficiently identified the optimal configuration for dementia classification.

Previous studies have relied largely on structured data or keyword-based extraction, both limited by documentation practices and input constraints. For example, Ernceoff et al. used ICD dementia- and cognition-related ICD codes to identify late-stage dementia, achieving modest performance (PPV=76.3%).^7^ Jaakkimainen et al. combined ICD codes and medication prescriptions to detect family physician-diagnosed early-onset dementia,^8, 9^ while Harding et al. integrated diagnosis codes, pharmacy codes, and specialty care visits, finding that at least two dementia diagnoses within 12 months yielded the best accuracy.^10^ Despite these efforts, all emphasized the need to incorporate free-text data to improve case detection. Recent studies have explored advanced approaches using NLP and machine learning. Ford et al. applied machine learning to Read codes, achieving an AUROC of 0.74 for undiagnosed dementia.^11^ Shao et al. combined structured and unstructured EHR data from the Veterans Health Administration to identify probable dementia cases, achieving a balanced sensitivity and specificity (∼83%).^13, 14^ These advances highlight the growing sophistication in dementia ascertainment. Unlike machine learning models, the RAG pipeline does not require large training datasets. Contrary to prior findings that incorporating free-text notes into structured EHR data improved performance,^11–14^ our study found that adding structured data alongside free-text notes in the LLM-based pipeline reduced accuracy. That said, structured data remain valuable for patients without available clinical notes.

A key strength of the RAG-based approach is its generalizability. It operates in a largely zero-shot manner and does not require prior knowledge, such as disease-specific keywords or ICD codes. Except for the prompt design, which was disease-specific, its core components, including document embedding, semantic retrieval, and response generation, are generalizable across disease types. This framework has broad potential to enhance disease ascertainment and phenotyping across conditions where critical information resides in unstructured notes.

Our study has several limitations. First, the evaluation was based on 623 patients from a single healthcare system, which may limit generalizability. Second, we tested a limited set of commercial and open-source LLMs without fine-tuning. Although GPT-4o performed well, its commercial nature raises concerns about cost and scalability. On the other hand, open-source models require substantial computational resources and may not match its performance. Third, the reference standard relied on human chart review, which is subject to error, particularly for patients with incomplete or pre-Epic records. These labels reflect subject-matter expert interpretation of the same data used for algorithm evaluation. It is not able to identify undocumented dementia patients. Finally, the work was used a limited number of data sources, including structured and free-text notes; future studies should integrate broader data sources to enhance completeness and accuracy.

Several directions for future research emerge from our findings. First, exploring fine-tuned or domain-adapted open-source LLMs optimized for clinical text could lower costs and enhance transparency. Second, prompt engineering remains an area for improvement, including developing methods to systematically identify the most effective prompts for specific tasks.

Finally, extending beyond binary classification to detect dementia onset, severity, and disease trajectories will require temporally aware retrieval and reasoning.

In summary, accurate dementia ascertainment from EHRs is critical for advancing real-world ADRD research and person-centered care. Our RAG-based, GPT-4o–enabled pipeline demonstrated high accuracy in identifying dementia status from free-text clinical notes, substantially outperforming traditional rule-based algorithms. These findings suggest that with effective prompt engineering and minimal training data, this approach can not only help detect undiagnosed dementia cases but also distinguish patients with dementia-related ICD codes who do not have true dementia. Future work will focus on applying the pipeline for large-scale retrospective EHR screening to support real-world ADRD research and improve clinical care delivery.

## CONCLUSION

This study developed and evaluated an LLM-based pipeline that uses RAG to retrieve salient information from large EHR free-text corpora and an LLM to ascertain patient-level dementia status, outperforming ICD-based rule algorithms and keyword-filtered LLM baselines. Future work will focus on external validation across diverse health systems and integrating this pipeline into predictive modeling and other downstream clinical applications.

## Data Availability

The datasets analyzed during the current study are not publicly available due to the EHRs being under the protection of HIPAA law and relevant laws for human subject research.

## Code Availability

The code for this study is publicly available on GitHub (https://github.com/RoboticReaper/MGB-Research-Dementia-Ascertainment-From-EHR)

## Author Contributions

L.W. conceptualized and designed the study, secured funding, and provided overall direction.

L.W. and R.Y. acquired and preprocessed the data. L.W., B.L., R.Y., Y.C., G.A.M. established and refined the reference standards. L.W., and B.L. conducted experiments, analyzed and interpreted the data, and drafted the manuscript. L.W. and G.M. supervised the research. All authors contributed to the manuscript revision and approved the final version.

## Declaration of Generative AI and AI-Assisted Technologies in the Manuscript Preparation Process

The authors used a generative AI tool (GPT-5, OpenAI) to assist with language editing and improving clarity of the manuscript text. The tool was not used for data analysis, interpretation of results, generation of scientific content, or drawing conclusions. All AI-assisted text was reviewed, edited, and verified by the authors, who take full responsibility for the integrity and accuracy of the manuscript.

