## Supplemental materials for "A retrieval-augmented generation large language model framework for accurate dementia identification from electronic health records"

**Table S1.** Performance of keyword-filtered large language model (LLM) classification pipelines.
**Table S2.** Prompts used in large language model (LLM)-based approaches for dementia classification.
**Table S3.** Detailed comparison of various settings in a retrieval-augmented generation (RAG)-based large language model (LLM) framework.
**Table S4.** Performance of retrieval-augment generation (RAG)-based large language model (LLM) pipelines.
**Table S5.** ICD codes, medications, cognitive assessments, and keywords for ADRD patient identification.
**Table S6**. Qualifying ICD-9-CM and ICD-10-CM codes for dementia.
**Table S7.** Keywords used for information filtering and note preparation for LLM.
**Table S8**. Template for aggregating relevant information from patient demographics, diagnosis, problem list and clinical notes.
**Table S9**. Large Language Models' Information.
**Table S10.** Embedding Models.

**Table S1.** Performance of keyword-filtered large language model (LLM) classification pipelines.

| Pipeline | Sensitivity (95% CI) | Specificity (95% CI) | PPV (95% CI) | NPV (95% CI) | Accuracy (95% CI) | F1 Score |
|---|---|---|---|---|---|---|
| **GPT-4o-P1** | **91.7 (87.6-95.9)** | **95.6 (93.7-97.5)** | **88.6 (83.9-93.3)** | **96.9 (95.3-98.5)** | **94.5 (92.8-96.3)** | **0.901** |
| **GPT-4o-P2** | 90.5 (86.1-94.9) | 96.0 (94.2-97.8) | 89.5 (84.9-94.1) | 96.5 (94.8-98.2) | 94.5 (92.8-96.3) | 0.900 |
| **GPT-4o-P3** | 91.7 (87.6-95.9) | 95.2 (93.2-97.1) | 87.6 (82.7-92.4) | 96.9 (95.2-98.5) | 94.2 (92.4-96.1) | 0.896 |
| **DeepSeek R1-P1** | 26.5 (19.8-33.2) | 98.9 (97.9-99.9) | 89.8 (81.3-98.3) | 78.3 (74.8-81.7) | 79.2 (76.0-82.4) | 0.409 |
| **Llama 3.3-P1** | 29.6 (22.7-36.5) | 98.7 (97.6-99.7) | 89.3 (81.2-97.4) | 79.0 (75.6-82.3) | 79.9 (76.8-83.1) | 0.444 |
| **Gemma 3-P1** | 99.4 (98.3-100.0) | 26.0 (22.0-30.0) | 33.3 (29.2-37.4) | 99.2 (97.5-100.0) | 45.9 (42.0-49.8) | 0.499 |

Abbreviations: PPV, positive predictive value; CI, confidence interval; NPV, negative predictive value

**Table S2.** Prompts used in large language model (LLM)-based approaches for dementia classification.

| Approach | Description |
|---|---|
| **System prompt** | **system_prompt**='''You are a knowledgeable and elite professional healthcare specialist, able to answer complicated and medical related questions from clinical notes. When answering questions, answer based on your knowledge and the provided patient notes. The question will be answered by yes or no format. After answering the question, give thorough explanations and reasons for your decision. Include specific information from the notes and cite the report number when it's helpful. Make your answer evidence based. Start your response with either YES or NO, fall back to NO if unsure. Proceed with explanations AFTER you've given your answer. Example: "Yes, the patient has dementia..." or "No, the patient does not have dementia", but NEVER start with "Based on the provided documentation"... because the response did not immediately start with the answer.''' |
| **User Prompt** | {**Patient_Data**}<br>Given information above, answer the following question: {**LLM_Query**} |
| **LLM_Query_P1** | Does the patient have dementia? Determine if there is an explicit and confirmed diagnosis of dementia specifically for this patient (not a family member or relative). Disregard mentions of cognitive complaints, medications, treatment trials (e.g., rivastigmine or Aricept), or indirect assessments (such as work-ups or evaluations) that do not explicitly state a dementia diagnosis. Consider more recent notes with more importance than older ones, and allow newer notes to negate older diagnosis if dementia information is conflicting.<br><br>If results of cognitive tests like MMSE, MOCA, or Mini-cog are mentioned, interpret them as following:<br>MMSE: 24 and higher: Normal cognition, no dementia. 19-23: mild dementia. 10-18: moderate dementia. 9 and lower: severe dementia.<br><br>MOCA: 26 or above: normal. 18-25: mild cognitive impairment. 10-17: moderate cognitive impairment. Less than 10: severe cognitive impairment. |

| | |
|---|---|
| | Mini-cog: If 0 of out 3 words are recalled, it is a positive screen for dementia, regardless of the clock-drawing. If all 3 words are recalled, it is immediately a negative screen for dementia, regardless of the clock-drawing. If the patient recalls 1-2 words, then the clinician needs to further refer to the clock-drawing: If there is 1-2 words (of out 3) for recall, with a normal clock, it is a negative screen for dementia. If there is 1-2 words (of out 3) for recall, with an abnormal clock, it is a positive screen for dementia.<br><br>ONLY START YOUR RESPONSE WITH "YES" or "NO", fall back to NO if unsure. Proceed with explanations AFTER you've given your answer. Example: "Yes, the patient has dementia..." or "No, the patient does not have dementia.” |
| **LLM_Query _P2** | As of the most recent available documentation, does the patient have dementia?<br>Carefully review the timeline of clinical documentation. Determine whether there is an explicit and confirmed diagnosis of dementia specifically for this patient (not a family member or relative).<br>Pay close attention to when information was recorded — prioritize more recent notes and diagnoses, but consider earlier entries to evaluate progression over time.<br>Disregard:<br>- Mentions of cognitive complaints alone<br>- Medications or treatment trials (e.g., rivastigmine, Aricept) without a confirmed diagnosis<br>- Indirect assessments (e.g., "work-up for dementia" or "referred for evaluation") unless they result in a clear diagnosis<br><br>ONLY START YOUR RESPONSE WITH "YES" or "NO", fall back to NO if unsure. Proceed with explanations AFTER you've given your answer. Example: "Yes, the patient has dementia..." or "No, the patient does not have dementia", but NEVER start with "Based on the provided documentation"... because the response did not immediately start with the answer. |
| **LLM_Query _P3** | Step 1: Analyze the Timeline of Documentation<br>Review the complete timeline of clinical notes and diagnoses to determine if a diagnosis of dementia was explicitly made for this patient (not for a family member).<br>Prioritize more recent clinical notes and diagnoses, ensuring they align with and confirm earlier findings. If there is only one dementia-related diagnosis code (e.g., Alzheimer’s disease or related dementia), subsequent documentation must continue to support or confirm the diagnosis for it to be valid.<br><br>Step 2: Assess Supporting Evidence<br>Consider Cognitive Test Scores: Include scores from cognitive assessments (e.g., MoCA, MMSE). Low scores alone do not confirm dementia but, if consistently impaired over time or paired with a diagnosis, they strengthen the case.<br>Evaluate Functional Status: If the patient is described as functionally |

| | |
|---|---|
| | independent or improving and there is no corroborating evidence of progressive decline, carefully reassess the validity of the earlier dementia diagnosis.<br>Transitions in Diagnosis: If earlier records list dementia but newer documentation clarifies mild cognitive impairment (MCI) or indicates improvement, favor the most recent clinically-supported status only if it contradicts earlier findings.<br><br>Step 3: Confirm Continuity of Evidence<br>Explicitly check whether the latest clinical documentation reaffirms an earlier dementia-related diagnosis. If the most recent evidence does not explicitly confirm or support the earlier diagnosis, reassess its validity.<br><br>Step 4: Disregard Irrelevant Mentions<br>Ignore the following unless explicitly tied to a clinical confirmation of dementia:<br>Memory concerns or subjective complaints without diagnosis.<br>Medications (e.g., donepezil, rivastigmine) unless paired with a documented diagnosis.<br>Phrases like “rule out dementia,” “referred for cognitive workup,” or “possible Alzheimer’s” unless subsequent documentation confirms diagnosis.<br>Isolated billing codes or problem list entries unless supported by clinical notes or functional assessments.<br><br>Step 5: Answer Classification<br>Provide your answer based on the following criteria:<br><br>"YES" if:<br>There is an explicit dementia diagnosis in the most recent clinical documentation and it aligns with or confirms earlier findings.<br>The diagnosis is supported by clinical observations, cognitive testing, or evidence of progressive functional decline.<br>"NO" if:<br>There is no confirmed dementia diagnosis in the most recent documentation.<br>The latest evidence indicates MCI or contradicts earlier mentions of dementia.<br>The earlier dementia diagnosis is unsupported by subsequent clinical findings, particularly when there is only one dementia-related diagnosis code. |

Note: Patient_Data is the placeholder where patient data were inserted. LLM_Query is the placeholder where query was inserted.

**Table S3.** Detailed comparison of various settings in a retrieval-augmented generation (RAG)-based large language model (LLM) framework.

| Experiment | Embedding | | | RAG Query | | Retrieval | | Structured data | | LLM | | | | | Prompt | | |
|---|---|---|---|---|---|---|---|---|---|---|---|---|---|---|---|---|---|
| group | MPNet | Bio_ClinicalBERT | Large | Word | Sentence | MMR | Cosine | No | Yes | GPT-4o | Llama 3.3 | Gemma 3 | Med42 | DeepSeek-R1 | P1 | P2 | P3 |
| A1 | ☑ | | | ☑ | | ☑ | | ☑ | | ☑ | | | | | ☑ | | |
| A2 | | ☑ | | ☑ | | ☑ | | ☑ | | ☑ | | | | | ☑ | | |
| A3* | | | ☑ | ☑ | | ☑ | | ☑ | | ☑ | | | | | ☑ | | |
| B1* | | | ☑ | ☑ | | ☑ | | ☑ | | ☑ | | | | | ☑ | | |
| B2 | | | ☑ | | ☑ | ☑ | | ☑ | | ☑ | | | | | ☑ | | |
| C1* | | | ☑ | ☑ | | ☑ | | ☑ | | ☑ | | | | | ☑ | | |
| C2 | | | ☑ | ☑ | | | ☑ | ☑ | | ☑ | | | | | ☑ | | |
| D1* | | | ☑ | ☑ | | ☑ | | ☑ | | ☑ | | | | | ☑ | | |
| D2 | | | ☑ | ☑ | | ☑ | | | ☑ | ☑ | | | | | ☑ | | |
| E1* | | | ☑ | ☑ | | ☑ | | ☑ | | ☑ | | | | | ☑ | | |
| E2 | | | ☑ | ☑ | | ☑ | | ☑ | | | ☑ | | | | ☑ | | |
| E3 | | | ☑ | ☑ | | ☑ | | ☑ | | | | ☑ | | | ☑ | | |
| E4 | | | ☑ | ☑ | | ☑ | | ☑ | | | | | ☑ | | ☑ | | |
| E5 | | | ☑ | ☑ | | ☑ | | ☑ | | | | | | ☑ | ☑ | | |
| F1* | | | ☑ | ☑ | | ☑ | | ☑ | | ☑ | | | | | ☑ | | |
| F2 | | | ☑ | ☑ | | ☑ | | ☑ | | ☑ | | | | | | ☑ | |
| F3 | | | ☑ | ☑ | | ☑ | | ☑ | | ☑ | | | | | | | ☑ |

Note: Rows are labeled as configurations. Each letter group represents a set of comparisons.
Starred (*) and rows indicate configurations that share identical components (embedding, retrieval, LLM, prompt). ☑ = included, empty = not used
Abbreviations: MPNet, all-mpnet-base-v2; Large, text-embedding-3-large; Cosine, cosine similarity; MMR, maximum marginal relevance; Word, "Dementia"; Sentence, "Does this person have dementia?"; LLM, large language model

**Table S4.** Performance of retrieval-augment generation (RAG)-based large language model (LLM) pipelines.

| Pipeline | Sensitivity (95% CI) | Specificity (95% CI) | PPV (95% CI) | NPV (95% CI) | Accuracy (95% CI) | F1 Score |
|---|---|---|---|---|---|---|
| **A1** | 87.6 (82.6-92.6) | 98.5 (97.3-99.6) | 95.5 (92.2-98.8) | 95.5 (93.6-97.4) | 95.5 (93.9-97.1) | 0.914 |
| **A2** | 78.1 (71.9-84.3) | 99.8 (99.6-100.0) | 99.3 (97.8-100.0) | 92.5 (90.1-94.8) | 93.9 (92.0-95.8) | 0.874 |
| **A3=B1=C1 =D1=E1=F1** | 87.1 (82.1-92.2) | 99.3 (98.6-100.0) | 98.0 (95.8-100.0) | 95.4 (93.5-97.3) | 96.0 (94.5-97.5) | 0.923 |
| **B2** | 89.9 (85.4-94.5) | 98.2 (97.0-99.5) | 95.0 (91.6-98.4) | 96.3 (94.6-98.0) | 96.0 (94.5-97.5) | 0.924 |
| **C2** | 87.0 (81.9-92.1) | 98.7 (97.6-99.7) | 96.1 (93.0-99.2) | 95.3 (93.4-97.2) | 95.5 (93.9-97.1) | 0.913 |
| **D2** | 93.6 (89.9-97.2) | 95.6 (93.7-97.5) | 88.9 (84.3-93.5) | 97.5 (96.1-99.0) | 95.1 (93.4-96.8) | 0.912 |
| **E2** | 62.1 (54.8-69.4) | 99.8 (99.4-100.0) | 99.1 (97.2-100.0) | 87.6 (84.8-90.5) | 89.6 (87.2-92.0) | 0.764 |
| **E3** | 66.3 (59.1-73.4) | 99.3 (98.6-100.0) | 97.4 (94.5-100.0) | 88.8 (86.0-91.5) | 90.4 (88.1-92.7) | 0.789 |
| **E4** | 79.9 (73.8-85.9) | 98.5 (97.3-99.6) | 95.1 (91.5-98.6) | 92.9 (90.6-95.2) | 93.4 (91.5-95.4) | 0.868 |
| **E5** | 62.1 (54.8-69.4) | 99.1 (98.3-1.0) | 96.3 (92.8-99.9) | 87.6 (84.7-90.4) | 89.1 (86.6-91.5) | 0.755 |
| **F2** | 87.6 (82.6-92.6) | 99.1 (98.3-1.0) | 97.4 (94.8-99.9) | 95.5 (93.7-97.4) | 96.0 (94.5-97.5) | 0.922 |
| **F3** | **91.1 (86.8-95.4)** | **98.5 (97.3-99.6)** | **95.7 (92.5-98.8)** | **96.8 (95.1-98.4)** | **96.5 (95.0-97.9)** | **0.933** |

Abbreviations: PPV, positive predictive value; CI, confidence interval; NPV, negative predictive value

**Table S5.** ICD codes, medications, cognitive assessments, and keywords for ADRD patient identification.

| Category | Details |
|---|---|
| ***Structured data*** | |
| Billing codes for dementias | ***ICD9***: 290.x, 294.1, 294.2, 331.0, 331.1, 331.2, 331.82, 780.93<br>***ICD10***: F01, F02.8, F03.9, G30.0, G30.1, G30.8, G30.9, G31.0, G31.1, G31.83, G31.9, R41.1, R41.3 |
| Billing codes for mild cognitive impairment | ***ICD9***: 331.83<br>***ICD10**:* G31.84, F06.7 |
| Dementia medication | Donepezil, Galantamine, Rivastigmine, Memantine, Donepezil-Memantine, Aducanumab, Lecanemab |
| Cognitive Assessments | Mini-Mental State Examination (MMSE), Mini-Cog, General Practitioner Assessment of Cognition (GPCOG), Eight-item Informant Interview to Differentiate Aging and Dementia (AD8), Saint Louis University Mental Status (SLUMS), Montreal Cognitive Assessment (MoCA), Memory Impairment Screen, Blessed Dementia Scale (BDS), Clinical Dementia Rating (CDR) |
| ***Keywords used in unstructured data*** | |
| Disease name related | "Alzheimer's disease", "ADRD", "dementia", "demented", "Mild cognitive impairment", "MCI", "Lewy body disease", "LBD", "Neurocognitive disorder", "Amnesia", "Frontotemporal lobar degeneration", "Primary progressive aphasia" |
| Dementia medication related | Donepezil, Aricept, Galantamine, Reminyl, Razadyne, Rivastigmine, Exelon, Memantine, Namenda, Namzaric, Aducanumab, Aduhelm,<br>Lecanemab, Leqembi, Donanemab, Kisunla |
| Cognitive assessments related | "Mini-Cog", "Mini-Mental State Examination", "MMSE", "General Practitioner Assessment of Cognition", "GPCOG", "Eight-item Informant Interview to Differentiate Aging and Dementia", "AD8", "Saint Louis University Mental Status", "SLUMS", "Montreal Cognitive Assessment", "MoCA", "Memory Impairment Screen", "Blessed Dementia Scale", "BDS", "Clinical Dementia Rating", "CDR" |

Abbreviations: ICD = International Classification of Diseases; MMSE = Mini-Mental State Examination; MoCA = Montreal Cognitive Assessment; MCI = Mild Cognitive Impairment; BDS = Blessed Dementia Scale; CDR = Clinical Dementia Rating.

**Table S6**. Qualifying ICD-9-CM and ICD-10-CM codes for dementia.

| Version | ICD Code | Definition |
|---|---|---|
| ICD-9-CM | 290.x | Dementias (including senile dementia, presenile dementia, and vascular dementia) |
| | 294.1x | Dementia in conditions classified elsewhere |
| | 294.2x | Dementia, unspecified |
| | 331.0 | Alzheimer’s disease |
| | 331.1x | Frontotemporal dementia |
| | 331.82 | Dementia with Lewy bodies |
| ICD-10-CM | F01.x | Vascular dementia |
| | F02.x | Dementia in other diseases classified elsewhere (e.g., Parkinson's) |
| | F03.x | Unspecified dementia |
| | G30.x | Alzheimer's disease |
| | G31.0x | Frontotemporal dementia |
| | G31.83 | Dementia with Lewy bodies |

Note: “x” serves as a placeholder, indicating that any digit may be used in that position to specify more detailed subtypes or variations of the diagnosis.

**Table S7.** Keywords used for information filtering and note preparation for LLM

| Category | Keywords |
|---|---|
| Disease-Related Terms | Alzheimer, dementia, demented, mild cognitive impair, amnesia, amnestic, frontotemporal, neurocognit, lewy body, Aphasia, memory, **MCI**, **FTD**, **LBD** |
| Medication-Related Terms | Donepezil, Aricept, Galantamine, Reminyl, Razadyne, Rivastigmine Exelon, Memantine, Namenda, Namzaric, Aducanumab, Aduhelm, Lecanemab, Leqembi, Donanemab, Kisunla |
| Symptom/cognitive function-Related Terms | confus, amnesia, forget, forgot, word, disorientation, attention, problem solving, poor safety awareness, executive dysfunction, mood, speech, disorientation, neuropsych, visuospatial, recall, processing speed, verbal fluency, executive, encoding, naming, orientation |
| Cognitive Assessment Tools /Tests | Mini-Cog, Mini-Mental State Examination<br>MMSE, GPCOG, AD8<br>Montreal Cognitive Assessment, MoCA<br>Brief Dementia Screening, BDS<br>Clinical Dementia Rating, CDR<br>Saint Louis University Mental Status Exam, SLUMS<br>Clock |
| Normal Cognitive-Related Terms | alert, oriented |

*Abbreviations*: LLM, large language model; MCI, mild cognitive impairment; FTD, frontotemporal dementia; LBD, Lewy body dementia; MMSE, Mini-Mental State Examination; GPCOG, General Practitioner Assessment of Cognition; AD8, Alzheimer’s Disease 8 (a brief screening tool for dementia); MoCA, Montreal Cognitive Assessment; BDS, Blessed Dementia Scale; CDR, Clinical Dementia Rating; SLUMS, Saint Louis University Mental Status.
*Note*: keywords in bold were matched on an exact basis, typically for specific abbreviations or terms where partial matching might lead to unintended results. The rest keywords were matched based on partial text presence in a sentence.

**Table S8**. Template for aggregating relevant information from patient demographics, diagnosis, problem list and clinical notes.

| | |
|---|---|
| **Keyword-filtered LLM approach** | The patient was {**Age**} years old. On {**Date**}, the following were documented in the EHRs:<br>The patient was diagnosed with: {**Diagnosis**$_1$**, Diagnosis**$_2$**, …, Diagnosis**$_n$}<br>The problem list included: {**Problem**$_1$**, Problem**$_2$**, …, Problem**$_n$}<br>The clinical notes documented: {**Sentence**$_1$**, Sentence**$_2$**, …, Sentence**$_n$}. |
| **RAG-based LLM approach** | Below are some excerpts from clinical notes for this patient:<br><br>-----------------------------------------<br><br>Patient Notes:<br>{context}<br><br>-----------------------------------------<br><br>Problems List:<br>{problems}<br><br>Diagnoses List:<br>{diagnoses} |

**Table S9**. Large Language Models' Information

| Model | Parameter | Context length | Version |
|---|---|---|---|
| GPT-4o | Not Disclosed | 128k | 2024-11-20 |
| DeepSeek R1 | 70B | 131k | 2025-05-28 |
| Llama 3.3 | 70B | 131k | 2024-12-06 |
| Gemma 3 | 27B | 131k | 2025-03-12 |
| Med42 | 70B | 8k | 2024-06-27 |

**Table S10.** Embedding Models

| Model | Embedding dimension | Source |
|---|---|---|
| all-mpnet-base-v2 | 768 | SentenceTransformers |
| Bio_ClinicalBERT | 768 | Emily Alsentzer |
| text-embedding-3-large | 3072 | OpenAI |
